# Genomic predictors of response to antidepressants in Major depressive disorder (MDD): A GWAS-Based Study on Indian cohort

**DOI:** 10.1101/2024.11.25.24317869

**Authors:** Isha Goel, Gagan Hans, Nand Kumar, Subhradip Karmakar

## Abstract

**Background:** Major depressive disorder (MDD) affects a significant portion of the population, with approximately 70% of patients not achieving adequate remission using anti-depressant monotherapy, a condition known as inadequate treatment response (ITR). In the Indian population, few studies have explored the genetic associations of this response, and those that exist are underpowered. This study aims to identify single nucleotide variations associated with ITR in the Indian population. The goal is to develop a population-specific gene panel that can identify subjects at risk for ITR, allowing alternative treatment modalities.

**Methods:** Subjects satisfying inclusion-exclusion criteria were enrolled for the study following institutional ethical approval. We recruited 120 subjects with MDD and assessed their response to monotherapy (Escitalopram) using the Hamilton Depression Rating Scale (HAMD) before and 4-6 weeks after medication. Responders (n=45) and non-responders (n=75) were genotyped using the Infinium Global Screening SNP-Array-24 v3.0. A genome-wide association study (GWAS) was performed to identify SNPs associated with ITR.

**Results:** Twelve lead-significant SNPs with a threshold of < 1E-05 were identified, suggesting an association with ITR. Among these, four SNPs were located in the intronic/regulatory regions of the genes ***LRSAM1, EFCAB2, TRIM56*** and ***ZNF17***. The remaining eight SNPs were near genes *ALDH1A2, LIPC, MYOCD, SPRY2, ANKRD18B, CCDC54, TNS3, ANKRD46*, and *FCRL2*. These genes are involved in critical functions related to cell signalling, immune response, neurodevelopment, regulating intracellular levels, and transcription factor binding.

**Conclusions:** Our study identified several novel SNPs that may be associated with ITR in MDD patients, reported for the first time in an Indian cohort. Further investigations are underway to determine their clinical significance and potential to be used for screening individuals responding to drug interventions as user-friendly gene panels.

## 1. Introduction

Unipolar depression, also known as Major Depressive Disorder (MDD), is among the leading causes of disability worldwide, ranking fourth in terms of disease burden and accounting for 46 million disability-adjusted life-years (DALYs) in 2021. According to the Global Burden of Disease Study 2021, depression is the fifth leading cause of disability in India, as measured by DALYs. The prevalence of depressive disorders increased from 3,745 cases per million in 2019 to 4,418 cases per million in 2021, with Major Depressive Disorder (MDD) alone contributing to over 70% of these cases, affecting nearly 45.4 million individuals in 2021 (Global Burden of Disease Collaborative Network, 2024). MDD affects individuals across all genders, ages, and social backgrounds. MDD is characterized by persistent sadness, loss of interest in activities, and a range of emotional and physical issues, often leading to suicidal thoughts (WHO, 2023). Symptoms of MDD can vary widely among individuals. First-line antidepressant monotherapy prescribed includes Selective Serotonin Reuptake Inhibitors (SSRIs) such as citalopram and escitalopram, and Serotonin-Norepinephrine Reuptake Inhibitors (SNRIs) like duloxetine and venlafaxine (Johnson CF et al., 2022). Despite their widespread use, the chronic and episodic nature of MDD, combined with poor psychosocial functioning, significantly contributes to the global economic burden. While medications and psychotherapy are effective for many, approximately 30-50% of depression patients do not achieve sufficient symptom relief with monotherapy, a condition known as inadequate treatment response (ITR). The situation is particularly alarming in India, where more than 70% of patients experience inadequate treatment response (Rafeyan R., et al., 2020; Arvind BA., et al., 2019).

One of the critical challenges in managing MDD is its multifaceted nature, which arises from diverse causes, coupled with our limited understanding of its pathogenic mechanisms. Apart from environmental influences like stress and trauma, genomic factors play a crucial role in the development of depression. Research indicates that depression can be hereditary, with specific genes involved in neurotransmitter regulation, such as serotonin, linked to a higher risk (Sullivan et al., 2000). Additionally, genes related to neuroplasticity (e.g., *BDNF*), the immune system (e.g., *SPP1*), and the stress response (e.g., 5-*HTTLPR*) contribute significantly to the complexity of depression. (Sforzini, L., et al., 2024; Marlene et al., 2023; Caspi, A. et al., 2003). Variants in these genes can predispose individuals to depression and, importantly, influence how they respond to antidepressants. The National Institute of Mental Health (NIMH) conducted the sequenced Treatment Alternatives to Relieve Depression (STARD) study*, which identified genetic markers associated with the effectiveness of antidepressant treatments helping to tailor personalized therapeutic approaches (Laje G., et al., 2009). Table 1 provides a list of genetic variants that have been associated with the response to antidepressant therapy in STARD* study and few others (Francis J. et al., 2006; Lemonde S. et al., 2004; Hong CJ., et al., 2006; Villafuerte SM., et al., 2009; Kato M., et al., 2009; Paddock S, et al., 2007). The genetic variants identified in the STARD* study were primarily based on a population in the United States (Caucasian, Black, and Hispanic individuals), which may not fully represent the genetic diversity found in the Indian population. Due to significant genetic differences and unique genetic variants in the Indian population, the applicability of these findings may vary.

**Table 1:**
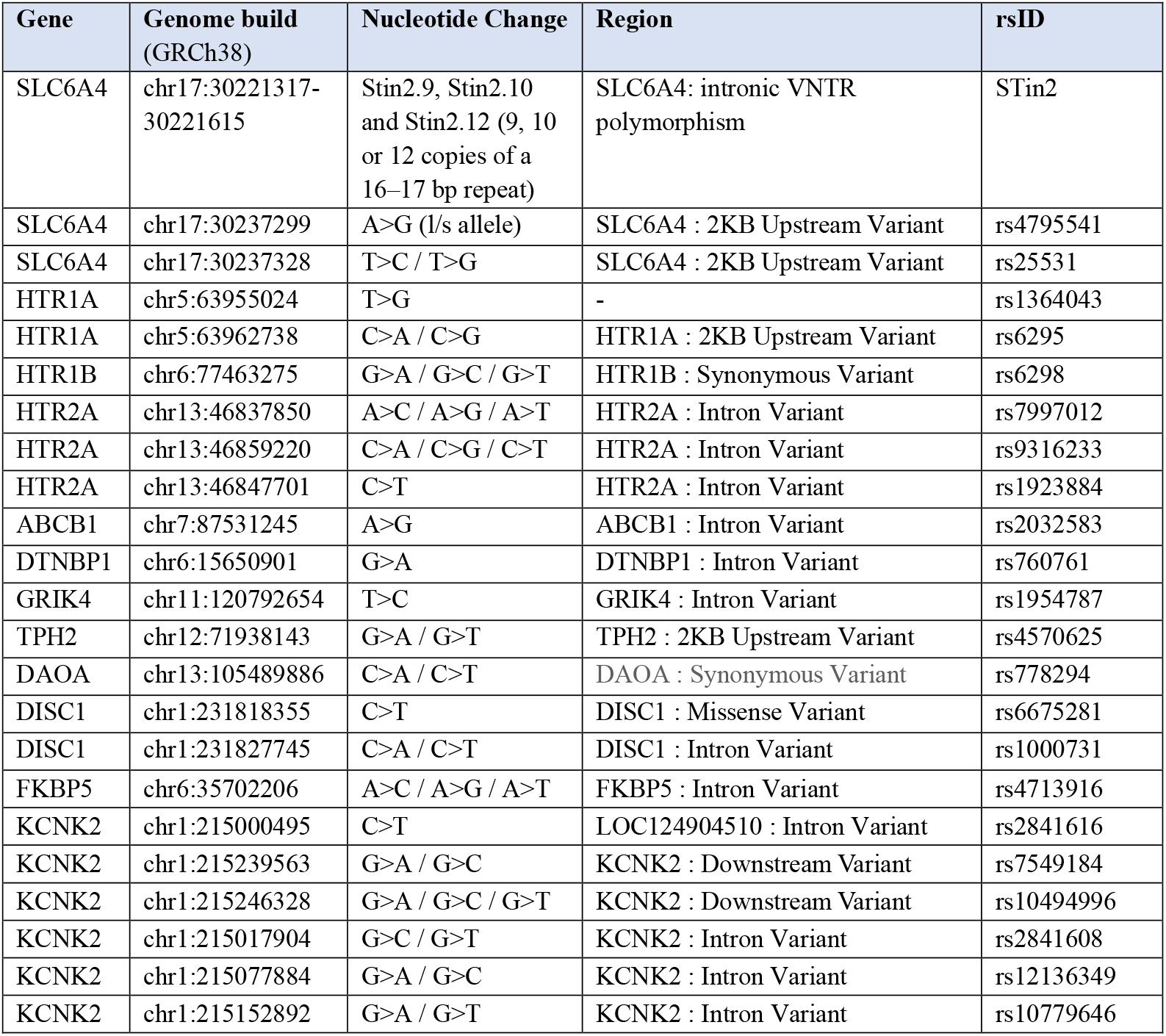
Table listing the details of the variants reported in literature to be associated with SSRI-treatment response.

| Gene | Genome build (GRCh38) | Nucleotide Change | Region | rsID |
| --- | --- | --- | --- | --- |
| SLC6A4 | chr17:30221317-30221615 | Stin2.9, Stin2.10 and Stin2.12 (9, 10 or 12 copies of a 16–17 bp repeat) | SLC6A4: intronic VNTR polymorphism | STin2 |
| SLC6A4 | chr17:30237299 | A>G (l/s allele) | SLC6A4 : 2KB Upstream Variant | rs4795541 |
| SLC6A4 | chr17:30237328 | T>C / T>G | SLC6A4 : 2KB Upstream Variant | rs25531 |
| HTR1A | chr5:63955024 | T>G | - | rs1364043 |
| HTR1A | chr5:63962738 | C>A / C>G | HTR1A : 2KB Upstream Variant | rs6295 |
| HTR1B | chr6:77463275 | G>A / G>C / G>T | HTR1B : Synonymous Variant | rs6298 |
| HTR2A | chr13:46837850 | A>C / A>G / A>T | HTR2A : Intron Variant | rs7997012 |
| HTR2A | chr13:46859220 | C>A / C>G / C>T | HTR2A : Intron Variant | rs9316233 |
| HTR2A | chr13:46847701 | C>T | HTR2A : Intron Variant | rs1923884 |
| ABCB1 | chr7:87531245 | A>G | ABCB1 : Intron Variant | rs2032583 |
| DTNBP1 | chr6:15650901 | G>A | DTNBP1 : Intron Variant | rs760761 |
| GRIK4 | chr11:120792654 | T>C | GRIK4 : Intron Variant | rs1954787 |
| TPH2 | chr12:71938143 | G>A / G>T | TPH2 : 2KB Upstream Variant | rs4570625 |
| DAOA | chr13:105489886 | C>A / C>T | DAOA : Synonymous Variant | rs778294 |
| DISC1 | chr1:231818355 | C>T | DISC1 : Missense Variant | rs6675281 |
| DISC1 | chr1:231827745 | C>A / C>T | DISC1 : Intron Variant | rs1000731 |
| FKBP5 | chr6:35702206 | A>C / A>G / A>T | FKBP5 : Intron Variant | rs4713916 |
| KCNK2 | chr1:215000495 | C>T | LOC124904510 : Intron Variant | rs2841616 |
| KCNK2 | chr1:215239563 | G>A / G>C | KCNK2 : Downstream Variant | rs7549184 |
| KCNK2 | chr1:215246328 | G>A / G>C / G>T | KCNK2 : Downstream Variant | rs10494996 |
| KCNK2 | chr1:215017904 | G>C / G>T | KCNK2 : Intron Variant | rs2841608 |
| KCNK2 | chr1:215077884 | G>A / G>C | KCNK2 : Intron Variant | rs12136349 |
| KCNK2 | chr1:215152892 | G>A / G>T | KCNK2 : Intron Variant | rs10779646 |

This study aims to address this critical issue within the Indian population, where genetic associations with treatment response have been minimally explored and existing studies lack sufficient power. By leveraging a genome-wide association study (GWAS), this research seeks to identify single nucleotide polymorphisms (SNPs) associated with ITR in Indian patients, paving the way for personalized treatment strategies. Building on this, our study will conduct a retrospective genotypic analysis of depressed patients treated at AIIMS clinics. By comparing patients requiring multi-drug therapy to those responding to monotherapy, we aim to validate reported proprietary markers and discover new ones, enhancing the genetic panel for predicting treatment response. This approach promises to improve clinical outcomes through earlier and more accurate identification of patients at risk for ITR, ultimately guiding more effective and individualized treatment plans.

## 2. Materials and methods

### 2.1 Subjects

The study was conducted at the Department of Biochemistry in collaboration with the Department of Psychiatry at the All-India Institute of Medical Sciences (AIIMS), New Delhi, India, from November 2022 to June 2024. Subjects presenting at the psychiatric outpatient department with symptoms of depressive disorder were assessed based on specific inclusion and exclusion criteria. Participants aged 18-65 years, who provided informed consent, and had non-psychotic major depressive disorder (DSM-V), HAMD scores ≥17, and no history of electroconvulsive therapy (ECT) were included. Those recommended for monotherapy (Escitalopram) treatment or already on multiple antidepressants due to non-response to escitalopram were further evaluated.

Exclusion criteria included major comorbidities such as cancer, surgery, heart disease, and known or suspected pregnancy during the study period. Subjects unwilling to provide consent were also excluded. Detailed patient information was recorded in both physical files and electronic documents, encompassing the patient information sheet, informed consent form, referral form with socio-demographic details, medical history, primary clinical diagnosis, and medication information. Depression severity was assessed using the 17-item HAMD scale through structured interviews.

To determine responders and non-responders to antidepressant treatment, the severity of patients’ depression was evaluated twice using the aforementioned scales at two time points. The first assessment was conducted on the day of patient enrolment, with those having a HAMD score ≥17 proceeding to further evaluation and blood collection (Evaluation I). Blood samples (4-5 ml) were drawn from consenting patients by an experienced practitioner and stored in sterilized EDTA vials. Patients were re-evaluated for depression severity after 4-6 weeks of antidepressant treatment using the HAMD scale (Evaluation II). A reduction in HAMD score of ≥50% categorized patients as responders, while those failing to meet this criterion were classified as non-responders. Patients already on multiple antidepressant therapies due to non-response to escitalopram were classified as non-responders to monotherapy.

### 2.2 Sample preparation

Blood samples collected at the OPD were immediately sent to the molecular lab in the Department of Biochemistry for DNA isolation. Using a genomic DNA extraction kit (Qiagen QIAamp Blood Midi-Kit – Catalogue no. 51185), genomic DNA (gDNA) was isolated from the blood following the standard kit protocol. The quality and quantity of the isolated gDNA were assessed using a spectrophotometer (Nano Drop Technologies, Wilmington, DE, USA) and gel electrophoresis. A DNA sample with a concentration of 100 ng/µL and an OD value (260/280 ratio) of approximately 1.8 was considered acceptable in terms of quality.

Subsequently, 2µg of purified gDNA was used for genotyping with the Illumina Infinium Global Screening BeadChip Array v3.0 (Illumina, San Diego, CA, USA). This array enables the genotyping of over 600,000 highly optimized multi-ethnic, curated clinical variants (based based on ClinVar, NHGRI-EBI and PharmGKB databases) spanning both coding and non-coding regions of the human genome. The genotyping data was generated in the form of .idat files, which were then analysed using Genome Studio 2.0.

### 2.3 Quality Control

Raw genotyping data files (.dat files) generated from the Illumina GSA bead-chip array were imported into GenomeStudio (v2.0) software (GenomeStudio Software (illumina.com)) using the Illumina hg38 manifest file and sample information sheet. SNP calls were generated and clustered, followed by quality control and filtering based on call rates, strand orientation, and gender estimation. Metadata was enriched with phenotypes of interest, and the cleaned data was exported into PLINK format for further processing. Using PLINK v1.9, the genotyping data underwent quality control procedures, including assessments of SNP and individual missingness, minor allele frequency, gender correction, heterozygosity, relatedness, and Hardy-Weinberg equilibrium (HWE) parameters. Highly heterozygous individuals, duplicates, closely related individuals, and SNPs in build-specific high linkage disequilibrium (LD) regions were removed.

### 2.4 Population Stratification and Imputation

The LD-pruned data was then checked for outliers using multidimensional scaling (MDS) analysis based on population stratification methodology, referencing population data from the 1000 Genomes Project phase 3. Outliers were identified using component-wise outlier detection and manual inspection of the MDS plot. Subjects that failed to follow population homogeneity were discarded. Phasing and imputation were performed using the Michigan Imputation Server 2 (Das S., et al., 2016) with the 1000 Genomes Project phase 3 dataset. This platform uses Eagle v2.4 to estimate haplotype phase and Minimac4 for imputation using a specific reference panel (HRC or 1000 Genomes). SNPs with R2 < 0.3 and minor allele frequency (MAF) < 0.05 were excluded.

### 2.5 Statistical Analysis and Annotation

The filtered SNPs were analysed for binary traits using the PLINK --assoc command, which employs a chi-squared test to identify SNPs significantly associated with the phenotype of interest. Quality and association results were visualized through QQ plots and Manhattan plots. Following this, SNP results were functionally annotated using Annovar to identify known and novel variants along with their associated genes.

## 3 Results

### 3.1 Patient enrolment

Total, 257 patients were screened at the psychiatric outpatient department of AIIMS. Out of these, 213 patients were enrolled in the study, with the remaining excluded due to age criteria, comorbidities, or refusal of consent. Within the enrolled group, 11 patients were excluded due to unsuccessful sample preparation. 82 patients dropped out during follow-up. Ultimately, 120 patients successfully completed all the assessments and medication. Table 2 shows demographic details of the patients enrolled and their response group based on the HAMD scale scores after successful evaluations.

**Table 2:** Demographic characteristics of the patients who have completed assessment.

| Characteristics | Patients N=120 |  |
| --- | --- | --- |
| Gender | Responders<br>N=45 | Non-responders<br>N=75 |
| Male (57) | 29 | 28 |
| Female (63) | 16 | 47 |
| Age |  |  |
| 18-40 years (81) | 31 | 50 |
| >40 years (39) | 14 | 25 |

Among the 120 patients studied, 37.5% were categorized as responders, whereas 62.5% were non-responders. Gender distribution showed that males constituted 47.5% of the total sample, with 24.2% of them being responders and 23.3% non-responders. Conversely, females made up 52.5% of the sample, with 13.3% responders and 39.2% non-responders, reflecting a higher prevalence of depression in females. Age-wise, 67.5% of the patients were between 18-40 years, with 25.8% responders and 41.7% non-responders, indicating a higher impact of depression in the younger age group. In the older age group (above 40 years), 32.5% of the patients were represented, with 11.7% being responders and 20.8% non-responders. These statistics suggest that depression disproportionately affects younger individuals more significantly.

### 3.2 QC and Population stratification

Genotyping calls from the raw data of all 120 samples (45 True-Responders, 65 Non-Responders, and 10 Resistant) were generated using Genome Studio 2.0. Out of the total 120 individuals, two were removed due to high heterozygosity, and one was removed due to duplicate sampling, resulting in 117 samples being retained for association analysis. SNP call rates for all individuals exceeded 98%. Of the initial 654,027 SNPs, 400,460 remained after filtering based on the aforementioned quality control metrics. Utilizing 1000g phase 3 population data, the remaining SNPs were subjected to MDS analysis for the assessment of population structure and identification of outliers. As shown in figure 3, all the 117 samples (labelled as OWN) from our study overlapped with individuals from SAS (South Asian) population that is composed of five sub-populations including Sri Lankan Tamils from the UK, Indian Telugu from the UK, Gujrati Indian from Houston, Texas, Punjabi from Lahore and Bengali from Bangladesh; the closest representative of Indian population in 1000 Genome phase 3 dataset. As inspected manually as well as computationally there were no significant outliers among the subjects under study, and all were processed further for imputation and association analysis.

**Figure 1.**
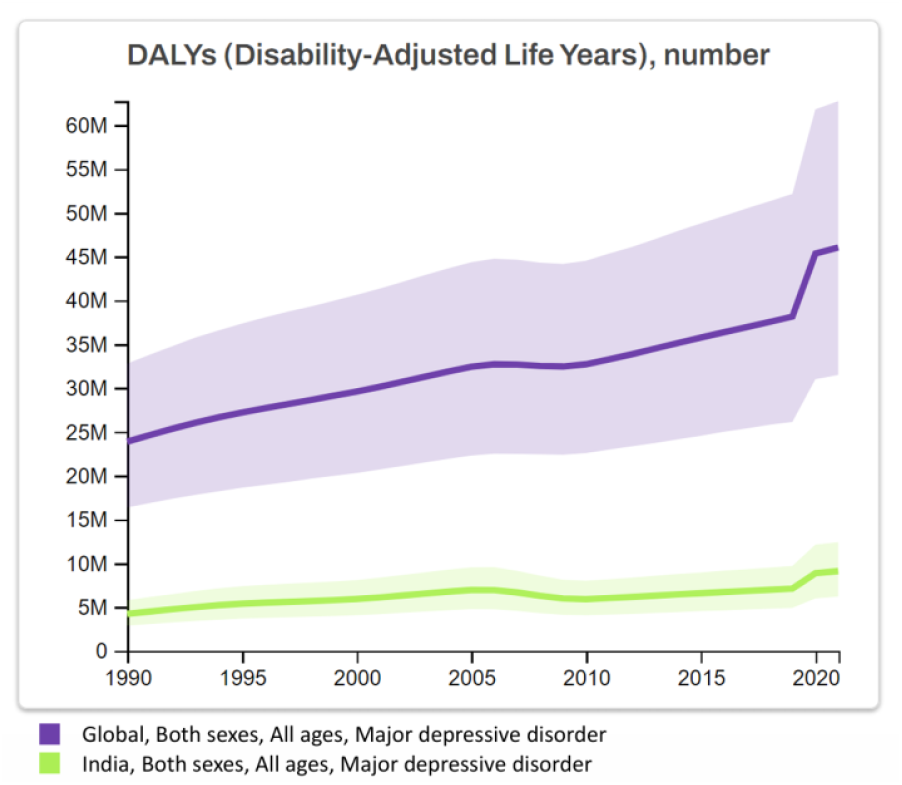
Graph showing the numbers of disability-adjusted life-years (DALYs) due to major depressive disorder in both the global and Indian contexts from 1990 to 2021. (Data and figure acquired from Global Burden of Disease Collaborative Network, 2024)

**Figure 2.**
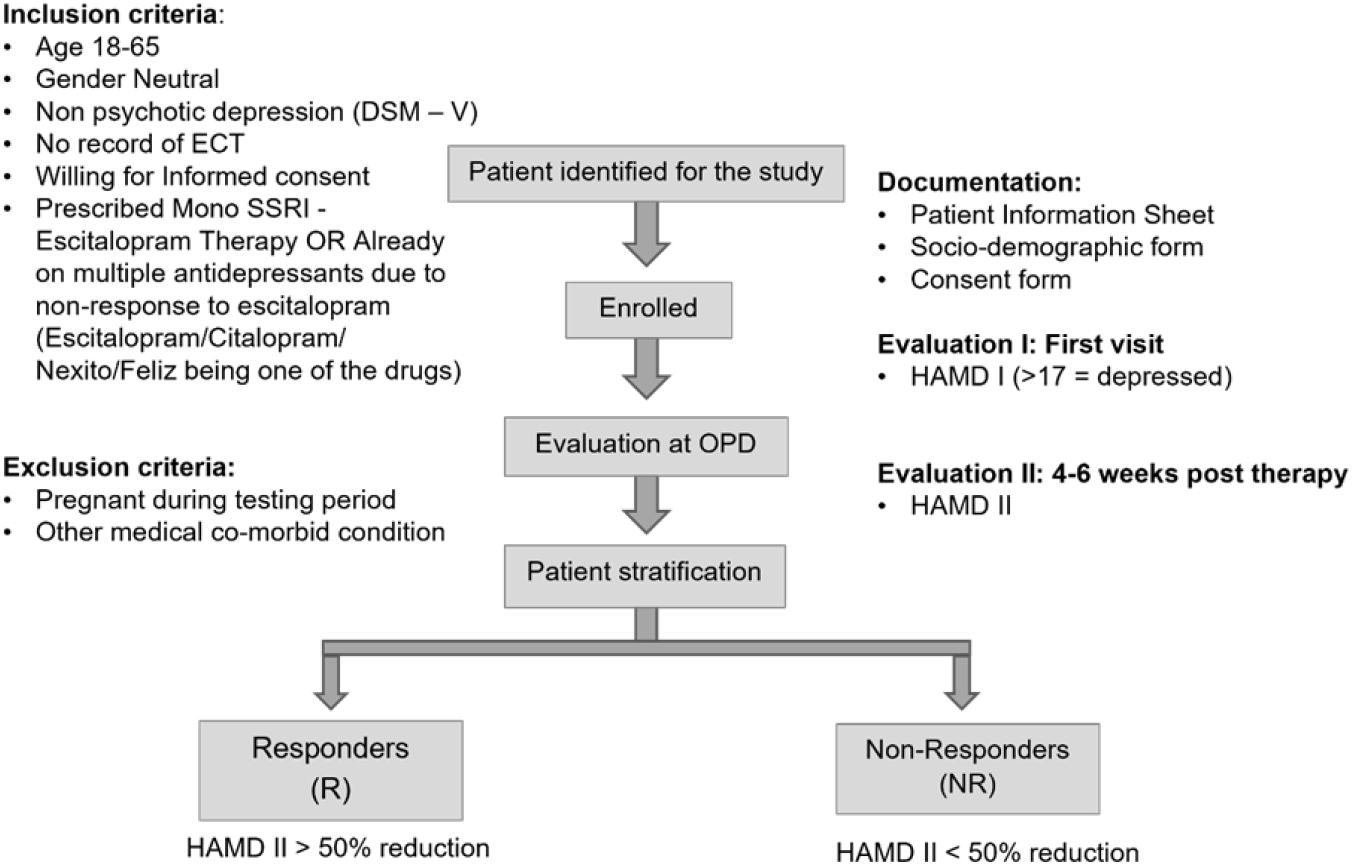
Work flow of sampling with details of Inclusion/Exclusion criteria and Evaluation metrics for the stratification of patients into responders and non-responder group.

**Figure 3.**
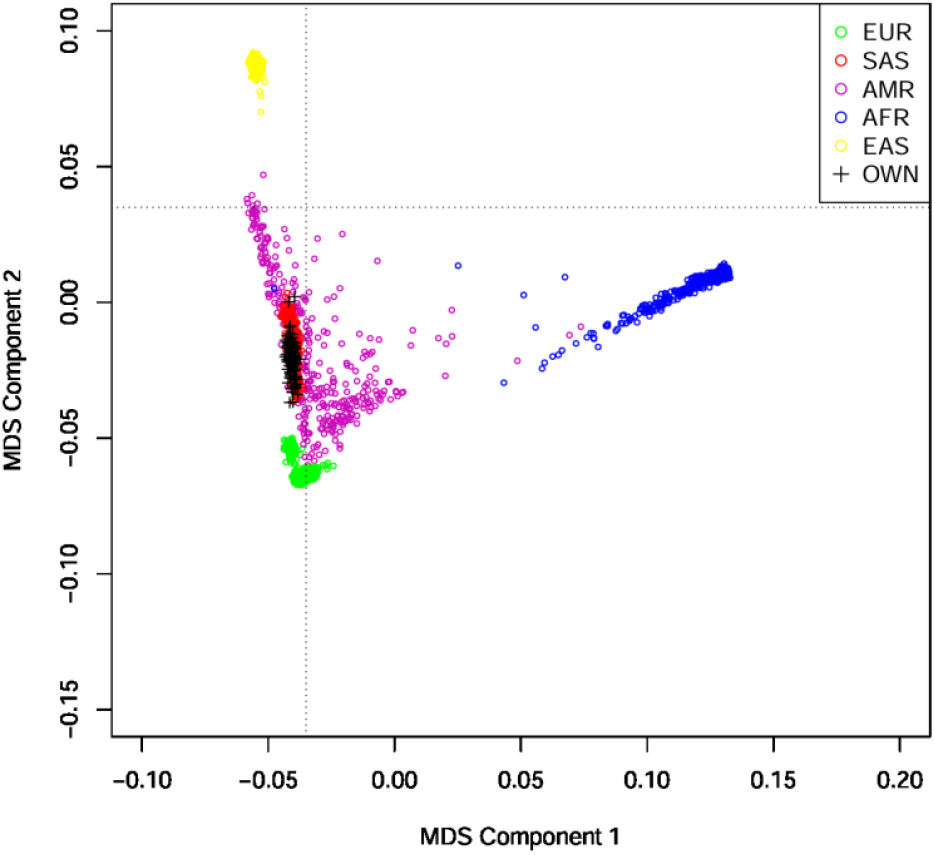
MDS plot showing the population structure of the 1000 Genome phase 3 data with individuals from different global populations including European (EUR), American (AMR), African (AFR), South Asian (SAS), and East Asian (EAS) along with the data of 117 individuals from this study labelled as OWN.

### 3.3 Phasing and Imputation

Additional pre-processing steps were performed for imputation as recommended on the Michigan Server of Imputation, following which 3,82,562 SNPs and 117 individuals passed all the filters and QC. These SNPs were then subjected to imputation using all the populations in the 1000 Genome phase 3 data. Variants with R2 score < 0.3 and MAF < 0.05 were filtered out, retaining 67,78,587 imputed SNPs with an average genotype calling rate of 1.0.

### 3.4 SNPs associated with ITR

Of the total 117 individuals, 73 belonged to the non-responder (non-responders to monotherapy + those on multi-antidepressant treatment due to non-response to monotherapy) groups and 44 belonged to the responder (responders to monotherapy) group. Association based on binary (Case/Control) trait was performed using --assoc option in PLINK v1.7. A Quantile-Quantile Plot was utilized to assess the quality of the data used for association based on Genomic inflation estimate: lambda (based on median chisq) = 1.0321, which was well within the acceptable range and represents good quality data, as shown in Figure 4. SNP results were represented in the form of a Manhattan plot (figure 5) and with a p-value cut-off of 1E-05 (1 x 10^-5^), a total of 78 significant variants including 12 lead SNPs were identified, suggestive of a significant association with ITR. Table 3-4 lists the details of the 12 lead SNPs identified suggestive of an association with ITR. Complete list of 78 significant variants is provided in the supplementary file S1.

**Table 3:** List of SNPs significantly associated with ITR in depression patients.

| Sno | CHR | POS | SNPID | P-value | Minor Allele | Major Allele | Frequency of Minor Allele in Cases | Frequency of Minor Allele in Controls | Odds Ratio | Asian Population Frequency |
| --- | --- | --- | --- | --- | --- | --- | --- | --- | --- | --- |
| 1 | 17 | 12569992 | 17:12569992:T:C | 2.96E-07 | C | T | 0.1575 | 0.4659 | 0.2144 | T=0.7148;<br>C=0.2852 |
| 2 | 13 | 80651301 | 13:80651301:G:A | 4.18E-07 | A | G | 0.2397 | 0.5682 | 0.2396 | G=0.66;<br>A=0.34 |
| 3 | 15 | 58359744 | 15:58359744:C:CACAA | 5.62E-07 | CACAA | C | 0.03425 | 0.25 | 0.1064 | ACA=1.00;<br>ACAAACA=0.00 |
| 4 | 9 | 33653459 | 9:33653459:G:A | 3.42E-06 | A | G | 0.08904 | 0.3295 | 0.1989 | G=0.85;<br>A=0.15 |
| 5 | 9 | 127457711 | 9:127457711:G:C | 4.50E-06 | G | C | 0.3288 | 0.6364 | 0.2799 | G=0.94;<br>C=0.06 |
| 6 | 3 | 107162120 | 3:107162120:G:A | 5.93E-06 | A | G | 0.2055 | 0.4886 | 0.2706 | G=0.64;<br>A=0.36 |
| 7 | 7 | 46940824 | 7:46940824:T:C | 6.33E-06 | C | T | 0.05479 | 0.2614 | 0.1638 | T=0.8554;<br>C=0.1446 |
| 8 | 1 | 245020277 | 1:245020277:C:G | 7.49E-06 | C | G | 0.06849 | 0.2841 | 0.1853 | C=0.15;<br>G=0.85 |
| 9 | 8 | 100440707 | 8:100440707:C:T | 8.30E-06 | T | C | 0.3699 | 0.6705 | 0.2885 | C=0.4960;<br>T=0.5040 |
| 10 | 7 | 101098969 | 7:101098969:G:A | 8.39E-06 | A | G | 0.5959 | 0.2955 | 3.516 | G=0.48;<br>A=0.52 |
| 11 | 19 | 57422202 | 19:57422202:A:G | 8.80E-06 | G | A | 0.3836 | 0.1136 | 4.853 | A=0.631;<br>G=0.369 |
| 12 | 1 | 157733448 | 1:157733448:T:A | 9.79E-06 | T | A | 0.3288 | 0.625 | 0.2939 | T=0.38;<br>A=0.62 |

**Table 4:** List of nearest genes of the SNPs significantly associated with ITR in depression patients.

| Sno | CHR | rsID | Type | Nearest Coding Gene | Nearest Coding Gene Name |
| --- | --- | --- | --- | --- | --- |
| 1 | 17 | rs4791503 | LINC00670 : Intron Variant | MYOCD | myocardin |
| 2 | 13 | rs1176317 | Intergenic | SPRY2 | sprouty RTK signaling antagonist 2 |
| 3 | 15 | rs1261660572 | Intergenic | ALDH1A2; LIPC | aldehyde dehydrogenase 1 family member A2; Lipase C |
| 4 | 9 | rs855549 | Intergenic | ANKRD18B | ankyrin repeat domain 18B |
| 5 | 9 | rs2243898 | LRSAM1 : Intron Variant | LRSAM1 | leucine rich repeat and sterile alpha motif containing 1 |
| 6 | 3 | rs116643249 | LINC00882(ncRNA) : Intron Variant | CCDC54 | coiled-coil domain containing 54 |
| 7 | 7 | rs11767105 | Intergenic | TNS3 | tensin 3 |
| 8 | 1 | rs901664 | EFCAB2 : Intron Variant | EFCAB2 | EF-hand calcium binding domain 2 |
| 9 | 8 | rs10107200 | LOC105375670 : Intron Variant | ANKRD46 | ankyrin repeat domain 46 |
| 10 | 7 | rs4489253 | TRIM56:1000B Downstream Variant | TRIM56 | tripartite motif containing 56 |
| 11 | 19 | rs10853894 | ZNF17 : 500B Downstream Variant | ZNF17 | zinc finger protein 17 |
| 12 | 1 | rs10430455 | Intergenic | FCRL2 | Fc receptor like 2 |

**Figure 4.**
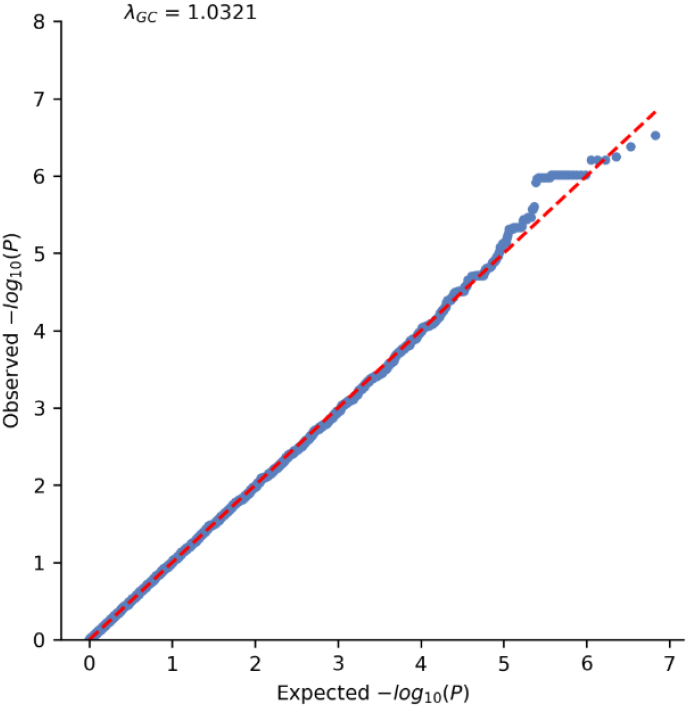
QQ (Quantile-Quantile) plot of Non-Responder vs Responder comparison showing the quality of the data with Genomic Inflation rate Estimate: Lambda = 1.0321, representing high-quality data.

**Figure 5.**
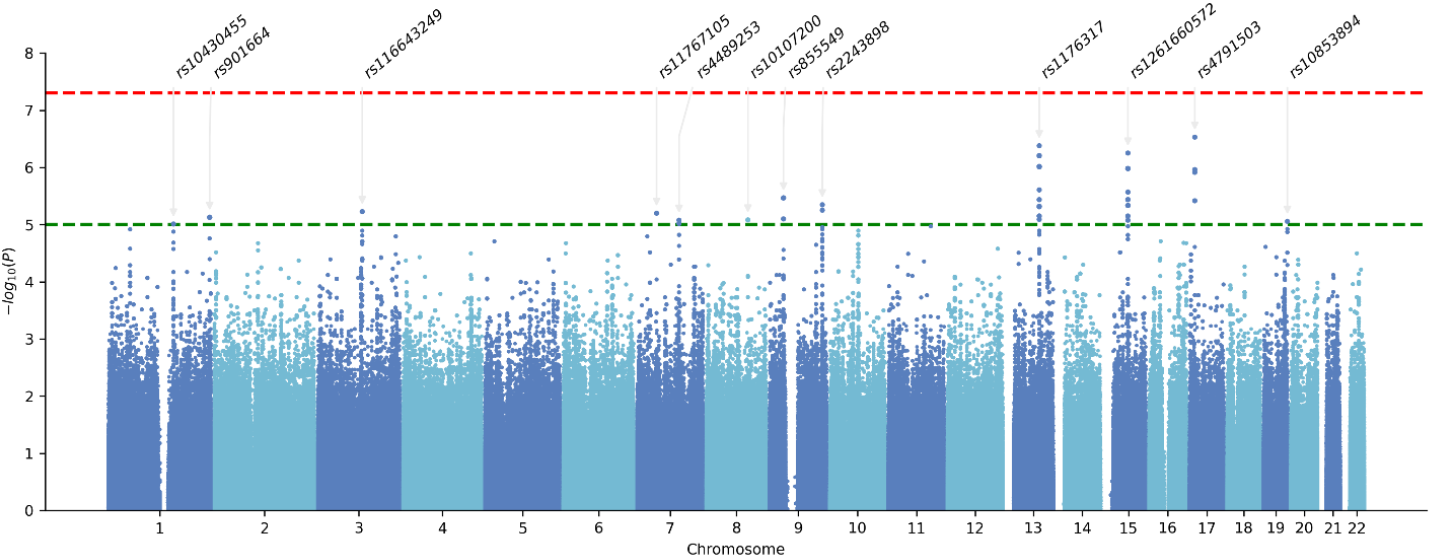
Manhattan plot demonstrating the relationship between SNPs and non-responders, highlighting some of the lead SNPs (rsIDs) with pval < 1 x 10^-5^. The green horizontal line at (P = 1E-05) represents the genome-wide significance criterion. Red line represents p-value cut-off of 5E-08.

The association analysis of single nucleotide polymorphisms (SNPs) identified several genetic variants significantly associated with antidepressant treatment response. Among the most notable findings, rs4791503 on chromosome 17 exhibited a highly significant p-value of 2.96×10^−7^. The minor allele (C) was observed at a frequency of 0.1575 in cases compared to 0.4659 in controls, resulting in an odds ratio of 0.2144. This SNP, an intron variant within the *LINC00670* gene, is situated near the *MYOCD* gene, which encodes myocardin, a critical regulator of smooth muscle gene expression. Another significant SNP, rs1176317 on chromosome 13, displayed a p-value of 4.18×10^−7^. Here, the minor allele (A) was present in 23.97% of cases and 56.82% of controls, yielding an odds ratio of 0.2396. This variant is adjacent to the *SPRY2* gene, known for its role in modulating receptor tyrosine kinase signaling pathways. The SNP rs1261660572 on chromosome 15, with a p-value of 5.62×10^−7^, showed the minor allele (CACAA) frequency to be 0.03425 in cases versus 0.25 in controls, leading to an odds ratio of 0.1064. This SNP is located upstream of the *ALDH1A2* gene, which is involved in retinoic acid biosynthesis.

Further significant associations were found with rs855549 on chromosome 9 (p-value 3.42×10^−6^), rs2243898 on chromosome 9 (p-value 4.50×10^−6^), and rs116643249 on chromosome 3 (p-value 5.93×10^−6^). These SNPs are linked to the *ANKRD18B, LRSAM1*, and *CCDC54* genes, respectively, all of which play essential roles in protein interactions and signalling pathways. Additionally, the SNPs rs4489253 on chromosome 7 (p-value 8.39×10^−6^) and rs10853894 on chromosome 19 (p-value 8.80×10^−6^) demonstrated the highest odds ratios of 3.516 and 4.853, respectively, indicating strong associations with the treatment response. These variants are situated near the *TRIM56* and *ZNF17* genes, involved in ubiquitination and transcription regulation, respectively. These findings underscore the potential genetic markers that can predict antidepressant treatment response. The identified genes are integral to various biological processes, including gene regulation, signal transduction, and cellular metabolism, thereby contributing to the observed variability in treatment response among individuals.

## 4 Discussion and limitations

Our study represents one of the pioneering GWAS investigations into inadequate drug response in depression within the Indian population, utilizing the Illumina Infinium SNP-array platform. The objective was to identify key genetic variations contributing to variability in treatment response. Subjects were carefully selected based on thorough clinical assessments.

Our study has certain limitations, primarily due to the moderate sample size, which is smaller than typical genome-wide association studies (GWAS). The vast genetic diversity within India’s large population adds complexity, as the diverse genetic backgrounds and ethnicities across different regions introduce significant variability not fully captured by the SAS sample in the 1000G population dataset. This variability makes it difficult to generalize our findings to the entire population and can introduce confounding factors, reducing the study’s power to detect significant associations. Additionally, cultural stigma around psychological disorders in India often leads to underreporting and a lack of formal diagnoses, affecting the quality and accuracy of the data. We also witnessed significant dropouts in follow-ups, which is typical in longitudinal studies. Despite these shortcomings, our results are intriguing and of clinical significance given the fact that a considerable number of people who die by suicide have a mental health issue. Identifying individuals at risk early on could avoid such disasters through proper counselling and other preventive measures.

Our study successfully identified several genetic variants associated with treatment response, underscoring the importance of including diverse populations in genetic research. To our knowledge, this study is one of the first to explore SNP-based heritability of inadequate drug response in depression among the Indian population. Most previous GWAS studies have focused on American ancestries. Our findings highlight the need for larger, more inclusive studies to understand the genetic underpinnings of depression and its treatment response in diverse populations. This research marks a significant step toward identifying genetic variations that influence drug response in depression, laying the groundwork for future research and the potential development of personalized treatment strategies that consider the genetic diversity of the Indian population.

In our study, we identified over 25 significant intergenic variants on chromosome 15, located near the regulatory regions of genes *ALDH1A2* and *LIPC. ALDH1A2* (Aldehyde Dehydrogenase 1 Family Member A2) encodes an enzyme involved in retinoate signalling, ESR-mediated signalling, and cofactor metabolism. *LIPC* (Lipase C, hepatic type) plays a crucial role in plasma lipoprotein assembly, remodelling, and cholesterol metabolism, with previous studies linking lipoprotein levels to depression and cognitive behaviour (Jia QF et al., 2020). Additionally, one significant variant on chromosome 9 was located in the intronic region of the gene *LRSAM1*, which encodes a ring finger protein involved in intracellular trafficking, antigen processing, signal transduction, cell adhesion, and the adaptive immune response. Mutations in *LRSAM1* have been linked to neurodegenerative diseases such as Parkinson’s disease (Mishra R et al., 2020).

We also discovered four significant intronic variants on chromosome 1 associated with the *EFCAB2* gene (EF-Hand Calcium Binding Domain 2), which is involved in calcium ion binding. Although its function is not yet fully determined, the GENDEP clinical trial suggests its role in neurodevelopment and cognitive function, particularly in axon cargo transport and neurotransmitter release (Uher, R., 2009; Ren H et al., 2018). Furthermore, a downstream variant on chromosome 19 (rs10853894) was significantly associated with the *ZNF17* gene, a DNA-binding transcription factor implicated in numerous essential cellular processes, including transcriptional regulation, ubiquitin-mediated protein degradation, signal transduction, actin targeting, DNA repair, and cell migration. ZNF proteins have been associated with various neuro-related diseases, including Alzheimer’s, schizophrenia, depression, anxiety, trauma, epilepsy, and autism (Bu S et al., 2021).

Other notable genes, such as *TRIM56, MYOCD, SPRY2, ANKRD18B, CCDC54, TNS3, ANKRD46, and FCRL2*, were also identified, highlighting their involvement in innate immune response, nervous system development, PDGFR-beta signalling pathway, FGFR2/3 signalling, and receptor tyrosine kinase signalling. These findings provide insights into the genetic underpinnings of drug response in depression, suggesting potential targets for personalized treatment strategies.

Notably, none of the identified SNPs, genes, or loci have been previously reported in earlier GWAS studies on depression. This discrepancy can be attributed to the fact that most GWAS research has predominantly focused on populations of American and Caucasian ancestry, with limited studies conducted on the Asian population, particularly the Indian demographic. Despite this, the major functional ontologies of the identified genes such as Immune response, cell signalling, metabolic pathways, and transcriptional activities were found to overlap significantly with those reported in previous research.

## 5 Acknowledgments

The authors thank the clinical staff (Psychologists) Praggapti Ghosh, Dr. Pratibha Lamba, Tanya Bhatia, Geetanjali Arora, and Hafsa Masroor for their contributions to the psychological assessments. We also acknowledge Saurabh Dubey for his technical support, Shubham Dhiryan and Komal Kashyap for data entry.

## 6. Author Contributions

Conceptualization, S.K., N.K. and G.H.; methodology, I.G., S.K., N.K. and G.H.; software, I.G and S.K.; formal analysis, I.G.; investigation, S.K., G.H.; data curation, I.G., G.H.; writing—original draft preparation I.G.; writing—review and editing, I.G., S.K., N.K. and G.H; supervision, S.K., N.K. and G.H; project administration, S.K., N.K. and G.H; funding acquisition, S.K., N.K.

## 7. Institutional Review Board Statement

Institute Ethics Committee of All India Institute of Medical Sciences, New Delhi gave ethical approval for this work, file no. IEC-523/05.06.2020, RP-16/2020

## 8. Informed Consent Statement

Informed consent was obtained from all subjects involved in the study.

## 9. Data Availability Statement

All data produced in the present study are available upon reasonable request to the authors.

## 10. Conflicts of Interest

The authors declare no conflict of interest.

## 11. Funding Statement

This research was funded by ICMR to vide project No. 5/4-4/187/M/2020-NCD-II

## Notes

### Competing Interest Statement

The authors have declared no competing interest.

### Summary of Updates

Figure 5 was cropped in the previous version and thus revised. Acknowledgments revised, Page no, line no. added.

